# Management and outcomes of myelomeningocele-associated hydrocephalus in low- and middle-income countries: a systematic review and meta-analysis protocol

**DOI:** 10.1101/2022.06.13.22276320

**Authors:** Berjo Dongmo Takoutsing, Alvaro Yanez Touzet, Jay J. Park, Seong Hoon Lee, Emily R. Bligh, Abdullah Egiz, Conor S. Gillespie, Anthony Figaji, the Neurology and Neurosurgery Interest Group

## Abstract

**Introduction:** Hydrocephalus and myelomeningocele (MMC) place disproportionate burdens of disease on low and middle-income countries (LMICs). MMC-associated hydrocephalus and its sequelae result in a spectrum of severely devastating clinical manifestations, for which LMICs are disproportionately unprepared in terms of human, capital, and technological resources. This study aims to review and compare the management and outcomes of infant MMC-associated hydrocephalus in LMICs and high-income countries.

**Methods and analysis:** This systematic review and meta-analysis will follow the PRISMA 2020 guidelines. The following databases will be searched without restrictions on language, publication date, or country of origin: EMBASE, MEDLINE, The Cochrane Library, Global Index Medicus, African Journals Online, and SciELO. All peer-reviewed studies of primary data reporting management and outcomes of infant MMC-associated hydrocephalus will be included. Where high-quality homogeneous studies exist, meta-analyses will be conducted to compare the management and outcomes of MMC-associated hydrocephalus across socioeconomic and geographic regions of the world. The primary outcome will be treatment failure of the first-line hydrocephalus treatment, which we defined operationally as the performance of a second intervention for the same reason as the first. Secondary outcomes include time to failure, rates of mortality, and postoperative complications.

**Ethics and dissemination:** Ethical approval was not applicable because this study does not involve human participants. Dissemination strategies will include publication in a peer-reviewed journal, oral and poster presentations at conferences, and an interactive web application to facilitate interaction with the findings and promote the discussion and sharing of findings on social media.

**ARTICLE SUMMARY:** *Strengths and limitations of this study:*

- This review focuses on multiple treatment modes of a well-defined disease population.
- Six electronic databases that are commonly used across both high- and low-income countries will be searched.
- No restrictions on language, location, or publication date were placed during screening.
- Unpublished studies will not be sought.
- The representativeness of the sample will rely on the quality of reporting of myelomeningocoele-associated hydrocephalus in the literature.
- Only one operational definition of treatment failure—‘the performance of a second intervention for the same reason as the first’—will be sought.
- An interactive web application dashboard will be developed to facilitate the transparent interaction with our methods and findings and promote scientific discussion and scrutiny.

## INTRODUCTION

Hydrocephalus places a disproportionate burden of disease on low- and middle-income countries (LMICs) (1, 2). It affects approximately 1 in 1,000 infants worldwide, but its incidence may exceed 200,000 cases per year in developing regions like sub-Saharan Africa (3). Myelomeningocele (i.e., meningomyelocele or open spina bifida; MMC) is a common and severe spinal aetiology and constitutes a significant proportion of this population (4-6). Its prevalence also varies by geography but approximates 113 cases per 100,000 births in LMICs (7, 8), reaching 77–610 and 700 cases per 100,000 births in South Africa and Nigeria, respectively. These disorders result in a spectrum of clinical manifestations, among which MMC-associated hydrocephalus is one of the most common and debilitating (4, 9, 10).

In LMICs, the incidence of MMC-associated hydrocephalus is high and can affect as many as 75% of cases (11, 12). Times to diagnoses and treatment are often delayed in these settings and, if treatment is not promptly initiated, most patients do not survive beyond infancy (4). Treatment may involve surgery, which is prone to significant morbidity and mortality, particularly in low-resource settings (1, 3). Intervening at the earliest stage with the best possible treatment remains, therefore, a crucial step in the management of MMC-associated hydrocephalus.

Ventriculoperitoneal (VP) shunts are the current standard of treatment in LMICs (1). Increasingly, patients are being treated with endoscopic third ventriculostomy (ETV) or combinations of ETV and choroid plexus cauterization (CPC), whose success rates and perceptions are variable (1, 3, 4, 9). Although ETV/CPCs were first developed and validated in LMICs as the primary management for MMC-associated hydrocephalus, there remains limited aggregated data regarding their outcomes in the very countries these procedures were developed in. As such, the practice and outcomes of these techniques in LMICs warrant systematic review and meta-analysis. Previous work has examined the management and outcomes of hydrocephalus in HICs and LMICs (13, 14) albeit in unstratified medical subject headings (MeSH) age groups and aetiologies.

## STUDY AIMS AND OBJECTIVES

### Aim

This study aims to review and compare the management and outcomes of MMC-associated hydrocephalus in infants across countries and treatment modes.

### Objectives

To review and compare:

1. The first- and second-line use of VP shunts, combinations of ETV and CPC, and conservative management;
2. The rates and times to treatment failure of VP shunts, combinations of ETV and CPC, and conservative management; and
3. Measures of mortality and postoperative complications of VP shunts, combinations of ETV and CPC, and conservative management.

### Review questions

Primary question:

1. What are the rates of failure of VP shunts, ETV, combinations of ETV and CPC, and conservative treatments in infant MMC-associated hydrocephalus? Secondary questions:
2. What are the most frequent first- and second-line treatments in the management of infant MMC-associated hydrocephalus?
3. What is the time to failure of VP shunts, ETV, combinations of ETV and CPC, and conservative treatments in infant MMC-associated hydrocephalus?
4. What are the mortality and postoperative complication rates of VP shunts, ETV, combinations of ETV and CPC, and conservative treatments in infant MMC-associated hydrocephalus?
5. Are the management and outcomes of the MMC associated with the failure of the hydrocephalus treatment?

Subgroup analyses by country and treatment mode are planned (further details in **Methods** section). The primary outcome will be the rate of treatment failure of the first-line hydrocephalus treatment. Secondary outcomes include: time to failure, rates of mortality, and postoperative complications. Operational definitions of outcomes are provided in the **Methods**.

Due to its historical standing as the local standard of treatment, our principal hypothesis is that:

H_1_: VP shunts have the lowest rates of treatment failure in the infant MMC-associated hydrocephalus population.

Secondary hypotheses include:

H_2_: VP shunts are the most frequently used first-line treatment and have the lowest mortality and complication rates.

H_3_: Resource constrained environments are associated with late/worse MMC presentation amongst infants

H_4_: Late/worse MMC presentation is associated with high MMC treatment failure, mortality, and complication rates.

H_5_: MMC complication rates are associated with hydrocephalus treatment failure, mortality, and complication rates.

## METHODS

This systematic review will be conducted following the guidelines outlined by the Preferred Reporting Items for Systematic Reviews and Meta-Analyses (PRISMA) 2020 statement (15).

### Search

A search strategy was developed to identify research articles on MMC-associated hydrocephalus. This was adapted from McCarthy et al., 2019, and consisted of synonyms of ‘hydrocephalus’ and ‘myelomeningocele’ (**Supplementary Table 1**). The search was run on the following electronic databases, from inception until 5 October 2021: EMBASE, MEDLINE, The Cochrane Library, Global Index Medicus, African Journals Online, and ScieLO. No restrictions on language, location, or publication date were placed, and unpublished studies will not be sought. Searches in all databases will be re-run prior to final analysis.

### Study selection

#### Types of studies

We will include articles published in peer-reviewed journals with any of the following designs: original research, trials, cross-sectional and cohort studies, multiple case reports (i.e., >1 case), and case series. Opinion pieces, comments, letters, guidelines, editorials, single-case reports, reviews, meta-analyses, and qualitative studies will be excluded, as well as articles published in non-peer-reviewed journals.

#### Country income level

We will include studies whose data were collected in low-, lower-middle-, upper-middle-, and high-income economies, according to the World Bank Country and Lending Groups (16). Studies from high-income economies will be used as comparators. Studies whose data cannot be traced to one particular country will be excluded.

#### Types of participants

All studies of infants aged 2 years or younger with a clinical or imaging diagnosis of MMC-associated hydrocephalus will be included. Non-infants (i.e., >2 years) will be excluded, as will infants with diagnoses of lipomyelomeningocoele, spina bifida occulta, or unspecified spina bifida, regardless of association with hydrocephalus.

#### Types of interventions

Studies of infants who have undergone either surgical or conservative treatment for hydrocephalus will be included. Surgical treatment includes VP shunting, ETV, combined ETV and CPC, combined VP shunting and ETV, and combined VP shunting, ETV, and CPC. We defined conservative treatment as non-surgical medical or pharmacological treatment. Studies of infants who underwent treatment for MMC, but not hydrocephalus, will be excluded, as will patients who received neither surgical nor conservative interventions.

#### Types of outcome measures

We will include studies of primary data reporting measures of treatment failure. In this review, treatment failure will be operationally defined as the performance of a second intervention for the same reason as the first. Studies reporting measures of mortality, morbidity, postoperative complications, and follow-up duration will also be included for secondary analysis. Post-operative complications will be defined as the unfavourable result of an intervention that did not result in treatment failure, and mortality as either ‘intra-operative’, if death occurred during a procedure, or ‘perioperative’, if it occurred within 30 days of surgery. For conservative treatments, mortality will encompass deaths that occurred within 30 days of administration.

### Outcomes

The primary outcome of our study will be the rate of treatment failure for the first-line hydrocephalus treatment. Secondary outcomes will include time to failure, rates of mortality, and postoperative complications.

### Study selection

Search results will be uploaded to Rayyan (https://www.rayyan.ai) to facilitate de-duplication and independent, blinded screening (17). First, titles and abstracts will be screened by two independent reviewers against the inclusion criteria, and the eligibility of the selected abstracts will be determined by reading the full texts. Unless otherwise stated, conflicts will be resolved through discussion and, where consensus cannot be achieved, through arbitration by the senior author (AF).

### Data extraction

Two reviewers will independently extract and check the data extracted from the included studies using a standardised extraction proforma in Google Sheets (**Supplementary Table 2**). Any disputes will be settled by a third reviewer (BDT, AYT, or AF). Data items will include information on study and sample characteristics; vertebral level of MMC and aetiology of hydrocephalus; clinical presentation and method of diagnosis; treatment mode, timing, and follow-up; and primary and secondary outcomes. If data is insufficiently provided in the full text, we will contact the corresponding author to request for the missing information and wait up to two months for a response. Following correspondence, all available data will be reported and studies with missing data will not be eligible for inclusion in the meta-analysis. All data will be recorded and stored in a spreadsheet.

### Risk of bias assessment

Included studies will be assessed for risk of bias by two independent reviewers of the extraction team. Randomised studies will be assessed using Version 2 of the Cochrane risk-of-bias tool for randomised trials (RoB 2) (18). Non-randomised studies will be assessed using the ROBINS I tool (19). Risk of bias assessment will be used during the analysis and results presented in terms of the primary and secondary objectives of this study.

### Data analysis

Following extraction, data will be transferred to Python for analysis. Screening results will be presented using a PRISMA flow diagram. Study and sample characteristics will then be summarised using descriptive statistics; these may include: vertebral level of MMC; aetiology of hydrocephalus; method of diagnosis; treatment mode; indication for surgery (if surgical management); timing, and follow-up; and primary and secondary outcomes.

The use, failure, and mortality measures of VP shunts, combinations of ETV and CPC, and conservative management will then be meta-analysed. Although meta-analyses are planned, these will only become apparent after data extraction is complete. Publication bias will be assessed through the use of funnel plots and Egger’s test (20). If asymmetry is found in the funnel plot, the Trim-and-Fill method will be used to account for any potential publication bias (21). The overall estimates and 95% confidence intervals will be obtained using random-effects models as per established methods (22). Cochran’s Q test (P < 0.10) and the I^2^ statistic will be used to assess heterogeneity among studies (23). Unless otherwise stated, the statistical significance will be set at P < 0.05, and comparisons to high-income economies will be drawn.

Mortality, failure, and complication rates will however vary with hydrocephalus aetiology, age, mode of treatment, characteristics of MMC, socioeconomic region and, crucially, duration of follow-up. Therefore, subgroup analyses will be sought, as appropriate, and particular care will be taken when interpreting failure between shunt placement and endoscopic treatments, owing to contrasting patterns of failure over the shorter and longer terms. A combination of dichotomous, proportion, continuous, O–E, and variance meta-analyses are planned for this purpose, as appropriate, depending on the data, and may include a potential subgroup analysis of the primary outcome into short-term rates and long-term rates. Recognising that case reports may present unusual and/or protracted outcomes, and randomised trials may assess active interventions improving outcomes, analyses by study design are also planned.

### Strength of body of evidence

The confidence in cumulative evidence included in this review will be assessed using the GRADE approach. Two independent reviewers will use the GRADEpro Guideline Development Tool (GRADEpro GDT) to assess the quality of outcomes of this study. A third reviewer (BDT, AYT, or JJP) will settle disagreements.

### Ethics and dissemination

Ethics approval for this study was not applicable because this study did not involve human participants. Dissemination strategies will include publication in a peer-reviewed journal, oral and poster presentations at conferences, and an interactive web application to facilitate interaction with the findings and promote the discussion and sharing of findings on social media.

### Patient and public involvement

None

## Data Availability

All data produced in the present work are contained in the manuscript

## ACKNOWLEDGEMENTS

We thank Moniba Korch, Brian Ou Yong Ming, and Safia Farsana Shabeer for their contributions during the development and piloting of the extraction proforma.

## AUTHOR CONTRIBUTIONS

BDT, AYT, and AF were responsible for conceiving the article. AF is the guarantor. BDT, AYT, JJP, and CG wrote the manuscript. AE, AF, BDT, AYT, CG, ERB, JJP, and SHL provided a critical appraisal of the manuscript. All authors critically revised and approved the final manuscript. BDT and AYT contributed equally and are joint first authors of the manuscript.

## ETHICS APPROVAL

Ethics approval for this study was not applicable because this study did not involve human participants.

## DATA AVAILABILITY STATEMENT

All data relevant to the study are included in the article or uploaded as supplementary information.

## REFERENCES

1. Muir RT, Wang S, Warf BC. Global surgery for pediatric hydrocephalus in the developing world: a review of the history, challenges, and future directions. Neurosurg Focus. 2016;41(5):E11.

2. Weiss HK, Garcia RM, Omiye JA, Vervoort D, Riestenberg R, Yerneni K, et al. A Systematic Review of Neurosurgical Care in Low-Income Countries. World Neurosurg X. 2020;5:100068.

3. Kahle KT, Kulkarni AV, Limbrick DD, Jr., Warf BC. Hydrocephalus in children. Lancet. 2016;387(10020):788–99.

4. McCarthy DJ, Sheinberg DL, Luther E, McCrea HJ. Myelomeningocele-associated hydrocephalus: nationwide analysis and systematic review. Neurosurg Focus. 2019;47(4):E5.

5. Oumer M, Taye M, Aragie H, Tazebew A. Prevalence of Spina Bifida among Newborns in Africa: A Systematic Review and Meta-Analysis. Scientifica (Cairo). 2020;2020:4273510.

6. Njamnshi AK, Djientcheu Vde P, Lekoubou A, Guemse M, Obama MT, Mbu R, et al. Neural tube defects are rare among black Americans but not in sub-Saharan black Africans: the case of Yaounde - Cameroon. J Neurol Sci. 2008;270(1-2):13–7.

7. Lo A, Polšek D, Sidhu S. Estimating the burden of neural tube defects in low- and middle-income countries. J Glob Health. 2014;4(1):010402.

8. Zaganjor I, Sekkarie A, Tsang BL, Williams J, Razzaghi H, Mulinare J, et al. Describing the Prevalence of Neural Tube Defects Worldwide: A Systematic Literature Review. PLoS One. 2016;11(4):e0151586.

9. Ntimbani J, Kelly A, Lekgwara P. Myelomeningocele - A literature review. Interdisciplinary Neurosurgery. 2020;19:100502.

10. de Paul Djientcheu V, Njamnshi AK, Wonkam A, Njiki J, Guemse M, Mbu R, et al. Management of neural tube defects in a Sub-Saharan African country: the situation in Yaounde, Cameroon. J Neurol Sci. 2008;275(1-2):29–32.

11. Dewan MC, Rattani A, Mekary R, Glancz LJ, Yunusa I, Baticulon RE, et al. Global hydrocephalus epidemiology and incidence: systematic review and meta-analysis. J Neurosurg. 2018:1–15.

12. Mnguni MN, Enicker BC, Madiba TE. A perspective in the management of myelomeningocoele in the KwaZulu-Natal Province of South Africa. Childs Nerv Syst. 2020;36(7):1521–7.

13. Ben-Israel D, Mann JA, Yang MMH, Isaacs AM, Cadieux M, Sader N, et al. Clinical outcomes in pediatric hydrocephalus patients treated with endoscopic third ventriculostomy and choroid plexus cauterization: a systematic review and meta-analysis. J Neurosurg Pediatr. 2022:1–13.

14. Pande A, Lamba N, Mammi M, Gebrehiwet P, Trenary A, Doucette J, et al. Endoscopic third ventriculostomy versus ventriculoperitoneal shunt in pediatric and adult population: a systematic review and meta-analysis. Neurosurg Rev. 2021;44(3):1227–41.

15. Page MJ, McKenzie JE, Bossuyt PM, Boutron I, Hoffmann TC, Mulrow CD, et al. The PRISMA 2020 statement: an updated guideline for reporting systematic reviews. Bmj. 2021;372:71.

16. World Bank Data Help Desk. World Bank Country and Lending Groups 2022 [Available from: https://datahelpdesk.worldbank.org/knowledgebase/articles/906519-world-bank-country-and-lending-groups.

17. Rayyan. Rayyan – Intelligent Systematic Review 2022 [Available from: https://www.rayyan.ai/.

18. Sterne JAC, Savović J, Page MJ, Elbers RG, Blencowe NS, Boutron I, et al. RoB 2: a revised tool for assessing risk of bias in randomised trials. Bmj. 2019;366:l4898.

19. Sterne JA, Hernán MA, Reeves BC, Savović J, Berkman ND, Viswanathan M, et al. ROBINS-I: a tool for assessing risk of bias in non-randomised studies of interventions. Bmj. 2016;355:i4919.

20. Egger M, Davey Smith G, Schneider M, Minder C. Bias in meta-analysis detected by a simple, graphical test. Bmj. 1997;315(7109):629–34.

21. Shi L, Lin L. The trim-and-fill method for publication bias: practical guidelines and recommendations based on a large database of meta-analyses. Medicine (Baltimore). 2019;98(23):e15987.

22. DerSimonian R, Laird N. Meta-analysis in clinical trials. Control Clin Trials. 1986;7(3):177–88.

23. Higgins JP, Thompson SG, Deeks JJ, Altman DG. Measuring inconsistency in meta-analyses. Bmj. 2003;327(7414):557–60.

