## Supplementary material for "Management and outcomes of myelomeningocele-associated hydrocephalus in low- and middle-income countries: a systematic review and meta-analysis protocol"

**SUPPLEMENTARY INFORMATION**

**Supplementary Table 1. Search terms for all databases**

**EMBASE**

| # | **Query** |
|---|---|
| 1 | exp hydrocephalus/ or hydrocephalus.mp. |
| 2 | (myelomeningocele or meningomyelocele).mp. or exp meningomyelocele/ |
| 3 | 1 and 2 |
| 4 | limit 3 to human |
| 5 | limit 4 to (conference abstract or conference paper or “conference review” or editorial or erratum or letter) |
| 6 | 4 not 5 |

**MEDLINE**

| # | **Query** |
|---|---|
| 1 | exp Hydrocephalus/ or hydrocephalus.mp. |
| 2 | (myelomeningocele or meningomyelocele).mp. or exp Meningomyelocele/ |
| 3 | 1 and 2 |
| 4 | limit 3 to humans |
| 5 | limit 4 to (conference abstract or conference paper or "conference review" or editorial or erratum or letter) |
| 6 | 4 not 5 |

**PubMed**

| # | **Query** |
|---|---|
| 1 | (("meningomyelocele"[MeSH Terms] OR "meningomyelocele"[All Fields] OR "myelomeningocele"[All Fields] OR "myelomeningoceles"[All Fields] OR ("meningomyelocele"[MeSH Terms] OR "meningomyelocele"[All Fields] OR "meningomyeloceles"[All Fields])) AND ("hydrocephalus"[MeSH Terms] OR "hydrocephalus"[All Fields])) AND (humans[Filter]) NOT ((booksdocs[Filter] OR editorial[Filter] OR letter[Filter] OR meta-analysis[Filter] OR review[Filter] OR systematicreview[Filter])) |

NANSIG

### The Cochrane Library

| # | Query |
|---|---|
| 1 | hydrocephalus |
| 2 | MeSH descriptor: [Hydrocephalus] explode all trees |
| 3 | #1 or #2 |
| 4 | myelomeningocele or meningomyelocele |
| 5 | MeSH descriptor: [Meningomyelocele] explode all trees |
| 6 | #4 or #5 |
| 7 | #3 and #6 |
| 8 | #7 in Cochrane Reviews, Cochrane Protocols, Editorials |
| 9 | #7 not #8 |

### Global Index Medicus

| # | Query |
|---|---|
| 1 | (myelomeningocele or meningomyelocele) and hydrocephalus |

### African Journals Online

| # | Query |
|---|---|
| 1 | (myelomeningocele or meningomyelocele) and hydrocephalus |

### SciELO

| # | Query |
|---|---|
| 1 | (((myelomeningocele)) OR (meningomyelocele)) AND (hydrocephalus) |

**Supplementary Table 2. Data item descriptions**

| Data item | Field type | Description |
| --- | --- | --- |
| Study characteristics | | |
| Study ID | Dropdown | Unique ID associated with each included article |
| Title | Lookup | Article title, extracted from citation |
| Author | Lookup | Article author(s), extracted from citation |
| Year of publication | Date | Only year |
| Year of data collection | Date | Only year |
| Original language of article | Dropdown | |
| Country of the study | Dropdown | The country where the management was done, not the country where the first author is located. From World Bank Country and Lending Groups. If management was done in multiple countries, a separate entry will be provided for each country. |
| Income level of study setting | Lookup | Lookup in World Bank Country and Lending Groups using **Country of the Study** |
| Region of study setting | Lookup | Lookup in United Nations Geoscheme using **Country of the Study** |
| Study design | Dropdown | Select one from: RCT, Multiple case report, Case series, Cohort study, Cross sectional study, Retrospective study (unspecified), Prospective study (unspecified), or Other (Add as comment) |
| Number of infants with MMC-associated HC | Integer | This need not necessarily be the same as the sample size of the study (e.g., studies looking at non-MMC aetiologies of HC) |

<table>
<tr><td>Age<br>(Mean, median, standard deviation and/or range)</td><td>Integers</td><td>In days. If multiple measures of central tendency or deviation present, extract all. If there are subgroups whose ages are within two years of birth, extract data for those subgroups. Do not extract data for any subgroups outside our age range. Regex validation string for range only:<br>^(\d|\d\d|[1-6]\d\d|7[0-2]\d|73[0-1])\.?(\d|\d\d)?-(\d|\d\d|[1-6]\d\d|7[0-2]\d|73[0-1])\.?(\d|\d\d)?$</td></tr>
<tr><td>Sex<br>(Female, male)</td><td>Integer</td><td>Of Number of infants with MMCaH.</td></tr>
<tr><td colspan="3">MMC management and outcomes</td></tr>
<tr><td>Signs or symptoms of MMC</td><td>Validated free text</td><td>Of Number of infants with MMCaH. Regex validation string:<br>^(.)+=(\s)?(\d|\d\d|\d\d\d|\d\d\d\d)((,(.)+=(\s)?(\d|\d\d|\d\d\d|\d\d\d\d))+)?$</td></tr>
<tr><td>Vertebral level of MMC<br>(Cervical, thoracic, thoracolumbar, lumbar, lumbosacral, sacral)</td><td>Integer</td><td>Of Number of infants with MMCaH. Compute as raw count if not given as such (e.g., if given as a proportion or percentage). Do not double count infants across vertebral levels.</td></tr>
<tr><td>Timing of MMC closure</td><td>Dropdown</td><td>Select one from: Ante-natal or post-natal. If mixed cohort, a separate entry will be provided for each closure.</td></tr>
<tr><td>MMC closure procedure and number of patients</td><td>Validated free text</td><td>The name of the intervention and the number of patients who had that intervention. Regex validation string:<br>^(.)+=(\s)?(\d|\d\d|\d\d\d|\d\d\d\d)((,(.)+=(\s)?(\d|\d\d|\d\d\d|\d\d\d\d))+)?$</td></tr>
</table>

<table>
<tr><td>Timing of MMC closure after birth</td><td>Validated free text</td><td>Groups/subgroups as reported by the authors. Regex validation string: ^(.)+=(\s)?(\d|\d\d|\d\d\d|\d\d\d\d)((,(.)+=(\s)?(\d|\d\d|\d\d\d|\d\d\d\d))+)?$</td></tr>
<tr><td>Age at MMC closure<br>(Mean, median, standard deviation and/or range)</td><td>Integers</td><td>As in Age. Regex validation string for range only: ^(\d|\d\d|\d\d\d|\d\d\d\d|\d\d\d\d\d)\.?(\d|\d\d)?-(\d|\d\d|\d\d\d|\d\d\d\d|\d\d\d\d\d)\.?(\d|\d\d)?$</td></tr>
<tr><td>Intra-operative mortality of MMC closure</td><td>Integer</td><td>Death during procedure. Compute as raw count if not given as such (e.g., if given as a proportion or percentage).</td></tr>
<tr><td>Peri-operative mortality of MMC closure</td><td>Integer</td><td>Death within 30 days of a procedure. Compute as raw count if not given as such (e.g., if given as a proportion or percentage).</td></tr>
<tr><td>Post-operative complications of MMC closure</td><td>Integer</td><td>Raw counts for Wound breakdown, Wound infection, and/or Meningitis. Regex validation string for miscellaneous only: ^(.)+=(\s)?(\d|\d\d|\d\d\d|\d\d\d\d)((,(.)+=(\s)?(\d|\d\d|\d\d\d|\d\d\d\d))+)?$</td></tr>
<tr><td colspan="3">First line intervention for HC</td></tr>
<tr><td>Patient sample</td><td>Dropdown</td><td>Select from: Selected or Unselected.</td></tr>
<tr><td>Age at diagnosis of HC<br>(Pre-natal or post-natal)</td><td>Integers</td><td>As in Age.</td></tr>
<tr><td>Method of diagnosis of HC<br>(CT imaging, MRI imaging, Ultrasonography, Clinical manifestation only, and Miscellaneous)</td><td>Integers</td><td>Raw counts for number of infants diagnosed with these methods. Regex validation string for miscellaneous only: ^(.)+=(\s)?(\d|\d\d|\d\d\d|\d\d\d\d)((,(.)+=(\s)?(\d|\d\d|\d\d\d|\d\d\d\d))+)?$</td></tr>
</table>

<table>
<tr><td>Indications for HC surgery<br>(Full or bulging fontanelle, Increased head circumference, CSF leaking from the wound, Symptoms of raised pressure, Other indications)</td><td>Integers</td><td>Raw counts for number of infants with these indications for HC surgery. Symptoms of raised pressure may include: These include: prominent scalp veins, high pitched cry, irritability, seizure, poor feeding, developmental delay, vomiting, setting sun sign, head suture and diastasis. Regex validation string for other indications only:<br>^(.)+=(\s)?(\d|\d\d|\d\d\d|\d\d\d\d)((,(.)+=(\s)?(\d|\d\d|\d\d\d|\d\d\d\d))+)?$</td></tr>
<tr><td>Co-morbidities<br>(Congenital heart defects, Congenital gastrointestinal malformations, Congenital face and neck malformations, Miscellaneous)</td><td>Integers</td><td>Raw counts for number of infants with these co-morbidities. Co-morbidities may not be recorded here if they are consequences of MMC/HC. Regex validation string for miscellaneous only:<br>^(.)+=(\s)?(\d|\d\d|\d\d\d|\d\d\d\d)((,(.)+=(\s)?(\d|\d\d|\d\d\d|\d\d\d\d))+)?$</td></tr>
<tr><td>Exacerbating factors<br>(Meningitis, Ventriculitis or Any other brain infection)</td><td>Integers</td><td>Raw counts for number of infants who had either meningitis or ventriculitis or any other brain infection before the HC.</td></tr>
<tr><td>Temporising procedures<br>(Subgaleal shunting or EVD)</td><td>Integers</td><td>Raw counts for number of infants who underwent subgaleal shunting or EVD as temporising procedures.</td></tr>
<tr><td>1st-line intervention for HC<br>(Conservative, VP shunting, ETV, Combined ETV and CPC, Failed ETV that was converted to VP shunt, or Miscellaneous)</td><td>Integers</td><td>Raw counts for number of infants who underwent one of these procedures as first-line treatment for HC. Non-surgical treatment is interpreted as conservative. If the infant underwent both VP and ETV, determine which was the primary one. For an infant to be recorded as a failed ETV that was converted to VP shunt,</td></tr>
</table>

| | | |
|---|---|---|
| | | the procedures must have happened in one sitting; alternatively, determine which was the primary one. Regex validation string for **miscellaneous** only: ^(.)+=(\s)?(\d\|\d\d\|\d\d\d\|\d\d\d\d)((,(.)+=(\s)?(\d\|\d\d\|\d\d\d\|\d\d\d\d))+)?$ |
| Age at first-line procedure | Integers | As in **Age.** |
| Follow-up period | Integer | In weeks |
| Intra-operative mortality of 1st-line HC procedure | Integer | As in **Intra-operative mortality of MMC closure** |
| Peri-operative mortality of 1st-line HC procedure | Integer | As in **Peri-operative mortality of MMC closure** |
| Post-operative complications of 1st-line HC procedure (Wound, Procedure failure, Wound infection, Wound dehiscence, Meningitis, Ventriculitis, CSF leakage, Focal Neurological deficits, Overdrainage, Intracranial haemorrhage, Any biochemical disturbance, and Any other complications) | Integers | Raw counts for Wound, Procedure failure, Wound infection, Wound dehiscence, Meningitis, Ventriculitis, CSF leakage, Focal Neurological deficits, Overdrainage, Intracranial haemorrhage, Any biochemical disturbance, and Any other complications. Regex validation string for **any biochemical disturbance** and **any other complications**: ^(.)+=(\s)?(\d\|\d\d\|\d\d\d\|\d\d\d\d)((,(.)+=(\s)?(\d\|\d\d\|\d\d\d\|\d\d\d\d))+)?$ |
| Number of procedures per patient | Integers | Raw counts. |

| | | |
|---|---|---|
| (Mean, Median, Standard deviation, and/or Range) | | |
| Second line intervention for HC | | |
| Time to 1st procedure failure, if any (Mean, Median, Standard deviation and/or Range) | Integers | The time between 1st-line HC procedure and 2nd-line HC procedure. In weeks. |
| Any information on why the treatment failed | Free text | |
| 2nd-line HC procedure | Integers | As in **1st-line HC procedure** |
| Age at second-line procedure | Integers | As in **Age at first-line procedure;** except that in weeks. |
| Intra-operative mortality of 2nd-line HC procedure | Integer | As in **Intra-operative mortality of 1st line HC procedure** |
| Peri-operative mortality of 2nd-line HC procedure | Integer | As in **Peri-operative mortality of 1st line HC procedure** |
| Post-operative complications of 2nd-line HC procedure | Integers | As in **Post-operative complications of 1st-line HC procedure** |
| Any further procedures for HC (repeat data items in **Second line intervention for HC** subheading) | | |
| Overall outcomes | | |

| | | |
|---|---|---|
| Overall complication rate, if any (Mean, Median, Standard deviation, and/or Range) | Integers | Number of infants with any complications divided by the number of infants that underwent any surgery |
| Overall incidence rate, if any (Mean, Median, Standard deviation, and/or Range) | Integers | Number of procedure failures divided by the number of procedures. Also known as the overall failure rate |

CPC: Choroid plexus cauterisation, ETV: Endoscopic third ventriculostomy, EVD: External ventricular drainage, HC: Hydrocephalus, MMC: Myelomeningocele, VP: ventriculoperitoneal
